# How complete is the reporting of somatic sensory training interventions in individuals following a stroke? Protocol for a systematic review

**DOI:** 10.1101/2021.09.16.21263676

**Authors:** Daniel Feller, Caterina Pedri, Paolo Gozzer, Tiziano Innocenti, Francesca Trentin

## Abstract

**Background:** To implement intervention from research to clinical practice, treatments must be adequately described in randomized clinical trials. Specific reporting guidelines, such as the TIDieR checklist, have been developed to enhance the reporting of intervention in clinical trials.

**Objective:** We aim to evaluate the adherence to the TIDieR checklist in randomized controlled trials evaluating somatic sensory training interventions in individuals following a stroke.

**Material and Methods:** We will perform a systematic review of the literature, searching PubMed, CENTRAL, and PEDro for randomized controlled trials that evaluate the efficacy of any rehabilitative intervention on somatic sensation in patients with a history of stroke, independently form the comparator.

Two authors independently will evaluate the completeness of the reporting of the intervention using the TIDieR checklist.

A descriptive analysis of the total score and the individual items of the TIDieR will be produced.

**Ethics and dissemination:** A manuscript with results will be submitted for publication in a peer-reviewed journal in the rehabilitation field.

## BACKGROUND

Somatic sensory deficits are very prevalent and highly correlate with impaired quality of life in patients with a history of stroke. ^1^

Rehabilitation interventions, both active and passive, are the most frequently used treatments to prompt the recovery from sensory deficits. Importantly, to implement interventions from clinical trials to clinical practice, the treatment must be adequately described, particularly for complex interventions such as sensory retraining. Also, a proper treatment’s description is essential for the reproducibility of clinical trials and to perform systematic reviews of the literature. ^2^

The *“Template for Intervention Description and Replication”* (TIDieR) checklist, in addition to other reporting guidelines, has been developed to enhance the reporting of interventions in clinical trials.^3^ Despite that, numerous studies have demonstrated a lack of adequate reporting in exercise-based interventions. ^4 5^

We postulate that also sensory retraining interventions are poorly reported in primary studies. Therefore, we aim to evaluate the adherence to the TIDieR checklist in randomized controlled trials (RCTs) evaluating somatic sensory training interventions in individuals following a stroke.

## MATERIAL AND METHODS

### Study selection process

Primary studies will be searched in biomedical databases, grey literature (i.e., Google Scholar), and through citation strategies using the included studies’ reference list.

PubMed, the Cochrane Register of Controlled Trials (CENTRAL), and PEDro will be investigated without time restrictions until 1st September 2021. Supplementary material 1 reports the search strategy used in PubMed.

The literature search will be limited to articles written in English or Italian.

The study selection process will be performed independently by two researchers. Any disagreement will be resolved by consensus or by the arbitrary decision of a third author.

The online electronic systematic review software package (Rayyan QCRI) will be used to organize and track the selection process. ^6^

### Inclusion and exclusion criteria

We will include only RCTs that recruited adults (age > 18) with a history of a stroke with any sensory and/or motor deficit, independently from:

- Type of stroke
- Location of the lesion
- Stage of the stroke (e.g., acute and chronic)

We will focus on any type of rehabilitative intervention, including active and passive sensory training, applied to the upper/lower limb or trunk, delivered as stand-alone or adjunct to usual care.

Regarding the outcome, we will include trials with any clinical outcome (scales or tests) that measure somatic sensation. RCTs that uses cortical excitability and other neurophysiological measures as the only outcome will be excluded.

Scales that measure sensation in addition to other impairments will be included if a subscore for the sensation part is given (e.g., sensation sub score of the Fugl-Mayer Assessment).

### Data extraction

The data extraction process will be conducted independently by two reviewers. Any discrepancies will be resolved with a consensus between the two authors and eventually by a third author’s arbitrary decision.

The following data will be extracted:

- First author and year of publication
- Type of intervention
- Type of outcome of somatic sensation
- Declared use of the TIDieR (yes/no)
- Adherence to the TIDieR

The TIDierR checklist will be used to assess the completeness of the reporting of the intervention.

According to the explanation and elaboration statements, properly described items will be marked with “1”, incomplete or missing items with “0”, and not applicable items with “NA”. The rating will be based on the article’s full text and any mentioned information (e.g., study protocol and supplementary data files).

### Data synthesis

A summary table and narrative synthesis of extracted data of all included studies will be provided.

A descriptive analysis of the total score and the individual items of the TIDieR will be produced. “NA” items will be excluded from the analysis. Specifically:

- Total adherence to the TIDieR for every included study will be calculated (in percentage) as the total number of items reported out of the total number of applicable items
- Total adherence for each item of the TIDieR will be calculated (in percentage) as the number of studies that properly reported that item out of the total number of studies that have such item applicable

In addition, as a subgroup analysis, we will present the total adherence for each item of the TIDieR for every different type of intervention found in the literature (e.g., mirror therapy, neurostimulation).

## ETHICS AND DISSEMINATION

An adequate reporting of the interventions in RCTs is vital for many reasons: it enables the clinician to implement the treatment directly in clinical practice, permits the exact replication of the trial, and allows researchers to aggregate appropriately similar RCTs in meta-analysis.

To the best of our knowledge, no study has ever investigated the reporting of the intervention in RCTs evaluating the efficacy of sensory retraining in patients following a stroke. Therefore, this study will provide valuable information that may improve future research in stroke rehabilitation.

A manuscript with results will be submitted for publication in a peer-reviewed journal in the rehabilitation field.

## Supporting information

Supplemental material 1 - Search on PubMed

## Data Availability

Not applicable.

## Acknowledgments

Not applicable.

## Notes

### Competing Interest Statement

The authors have declared no competing interest.

### Funding Statement

None to declared.

