## Supplemental material 1 - Search on PubMed for "How complete is the reporting of somatic sensory training interventions in individuals following a stroke? Protocol for a systematic review"

**SUPPLEMENTARY MATERIAL 1** – Search strategy for PubMed

1. (cerebrovascular disorders OR stroke* OR cerebrovasc* OR apoplex* OR poststroke* OR hemipleg* OR hemipar* OR brain vasc* OR cerebral vasc*)
2. (somatosensory disorders OR sensation OR sensation* OR propriocept* OR kinesthe* OR touch* OR stereognosi* OR sensor* OR somatosensory* OR tactile OR thermal OR pinprick OR position sense)
3. (rehabilitation OR rehab* OR train* OR retrain* OR education OR re-education OR reeducation OR practice OR treatment* OR intervention*)
4. (randomized controlled trial[pt] OR controlled clinical trial[pt] OR randomized[tiab] OR placebo[tiab] OR clinical trials as topic[mesh:noexp] OR randomly[tiab] OR trial[ti] NOT (animals[mh] NOT humans [mh]))
5. #1 OR #2 OR #3 OR #4

**Complete search strategy:**

(cerebrovascular disorders OR stroke* OR cerebrovasc* OR apoplex* OR poststroke* OR hemipleg* OR hemipar* OR brain vasc* OR cerebral vasc*) AND (somatosensory disorders OR sensation OR sensation* OR propriocept* OR kinesthe* OR touch* OR stereognosi* OR sensor* OR somatosensory* OR tactile OR thermal OR pinprick OR position sense) AND (rehabilitation OR rehab* OR train* OR retrain* OR education OR re-education OR reeducation OR practice OR treatment* OR intervention*) AND (randomized controlled trial[pt] OR controlled clinical trial[pt] OR randomized[tiab] OR placebo[tiab] OR clinical trials as topic[mesh:noexp] OR randomly[tiab] OR trial[ti] NOT (animals[mh] NOT humans [mh]))
